# Humoral and cellular immune responses following BNT162b2 XBB.1.5 vaccination

**DOI:** 10.1101/2023.10.04.23296545

**Authors:** Metodi V. Stankov, Markus Hoffmann, Rodrigo Gutierrez Jauregui, Anne Cossmann, Gema Morillas Ramos, Theresa Graalmann, Michaela Friedrichsen, Inga Ravens, Tamara Ilievska, Jasmin Ristenpart, Anja Schimrock, Stefanie Willenzon, Gerrit Ahrenstorf, Torsten Witte, Reinhold Förster, Amy Kempf, Stefan Pöhlmann, Swantje I. Hammerschmidt, Dopfer-Jablonka Alexandra, Georg M. N. Behrens

## Abstract

SARS-CoV-2 Omicron XBB subvariants efficiently evade immunity from prior infection or vaccination, requiring vaccine adaptation. Here, we analyzed immunogenicity of an adapted vaccine, BNT162b2 Omicron XBB.1.5, which is currently used for booster vaccination. Booster vaccination significantly increased anti-Spike IgG, accompanied by expansion of cross-reactive memory B cells recognizing Wuhan and Omicron XBB.1.5 spike variants. Geometric mean neutralizing titers against XBB.1.5, XBB.1.16 and XBB.2.3, as well as cross-reactive responses against EG.5.1 and BA.2.86 increased significantly relative to pre-booster titers. Finally, the number of Wuhan and XBB.1.5 spike reactive IFN-γ-producing T cells significantly increased after booster vaccination. In summary, BNT162b2 Omicron XBB.1.5 vaccination resulted in potent neutralizing antibody responses against Omicron XBB variants, including the recent Omicron variants EG.5.1 (Eris) and BA.2.86 (Pirola), as well as XBB.1.5 reactive T cell responses, suggesting that booster vaccination will augment protection against these emerging variants.

## Main

Whilst the coronavirus disease 2019 (COVID-19) pandemic is transitioning into an endemic, the causative agent, severe acute respiratory syndrome coronavirus 2 (SARS-CoV-2), continues to circulate globally and to threaten public health. Omicron XBB sublineages, including the XBB.1.16 and EG.5.1 variants, are currently responsible for a large proportion of COVID-19 cases and the recent emergence of BA.2.86 is paralleled by an increase in cases in several countries. The BA.2.86 spike (S) protein contains roughly 60 mutations compared to the S protein of the virus circulating at the beginning of the pandemic (Extended Data Fig. 1), which are mostly located within the N-terminal domain (NTD) and the receptor binding domain (RBD), and result in robust evasion of neutralizing antibodies used for COVID-19 therapy or elicited upon infection or vaccination. Due to the efficient evasion of neutralizing antibodies of current XBB subvariants and BA.2.86, adapted vaccines are needed and monovalent Omicron XBB.1.5-containing vaccines have recently been approved and are now being rolled out. While preliminary evidence from the manufacturers suggests promising neutralization of emerging variants, real-world evidence is currently lacking^1^ and persisting immune imprinting^2,3^ might compromise elicitation of antibodies against new SARS-CoV-2 variants. Therefore, it is important to determine how novel mRNA vaccines impact immune surrogates of protection against most recent Omicron variants. In this analysis, we monitored immune responses in 65 participants from our ongoing COVID-19 Contact (CoCo) Study^4^ that were vaccinated with the updated monovalent BNT162b2 Omicron XBB.1.5/Raxtozinameran (30 µg) vaccine.

Most study participants had previously received a two-dose primary series, a third dose of the original mRNA (Wuhan, Wu) COVID-19 vaccine. In total, 66% had received 4 or more vaccinations before baseline and 30% of the participants had received a bivalent, adapted (Omicron BA.4/BA.5 + Wu strain) vaccine. Of all vaccinees, 84% reported a prior SARS-CoV-2 infection and 79% reported a SARS-CoV-2 infection during the period of Omicron dominance (since Jan 2022), with a mean of 1.1 Omicron antigen exposures. 9.5% reported neither a SARS-CoV-2 infection, nor an Omicron booster. The most recent SARS-CoV-2 infection occurred a median of 14.4 months ago, and no individual was excluded due to evidence of SARS-CoV-2 infection during the reported study period. The vaccination and blood collection schedule is detailed in the methods section, with additional demographic information provided in Extended Data Table 1.

After reaching a plateau in the post-vaccination increase of anti-Spike (S) IgG antibody in a subgroup of vaccinees on day eight (Online Methods and Extended Data Fig. 2), responses were assessed at a mean of 9.4 days post-vaccination in the entire cohort. Pre-vaccination, participants exhibited a mean of 1,422±1,153 BAU/mL (±SD) anti-spike (S) IgG antibodies and a mean of 199±140 RU/mL (±SD) Omicron IgG antibodies, which is within the range of responses after the first two BNT162b2 vaccinations ^4^ but below peak responses three weeks after the third vaccination^5^. Overall, Omicron XBB.1.5 booster vaccination resulted in a significant (p<0.001) 2.9 and 3.6-fold increase in anti-S IgG and Omicron anti-S IgG, respectively (Figure 1a).

**Figure 1:**
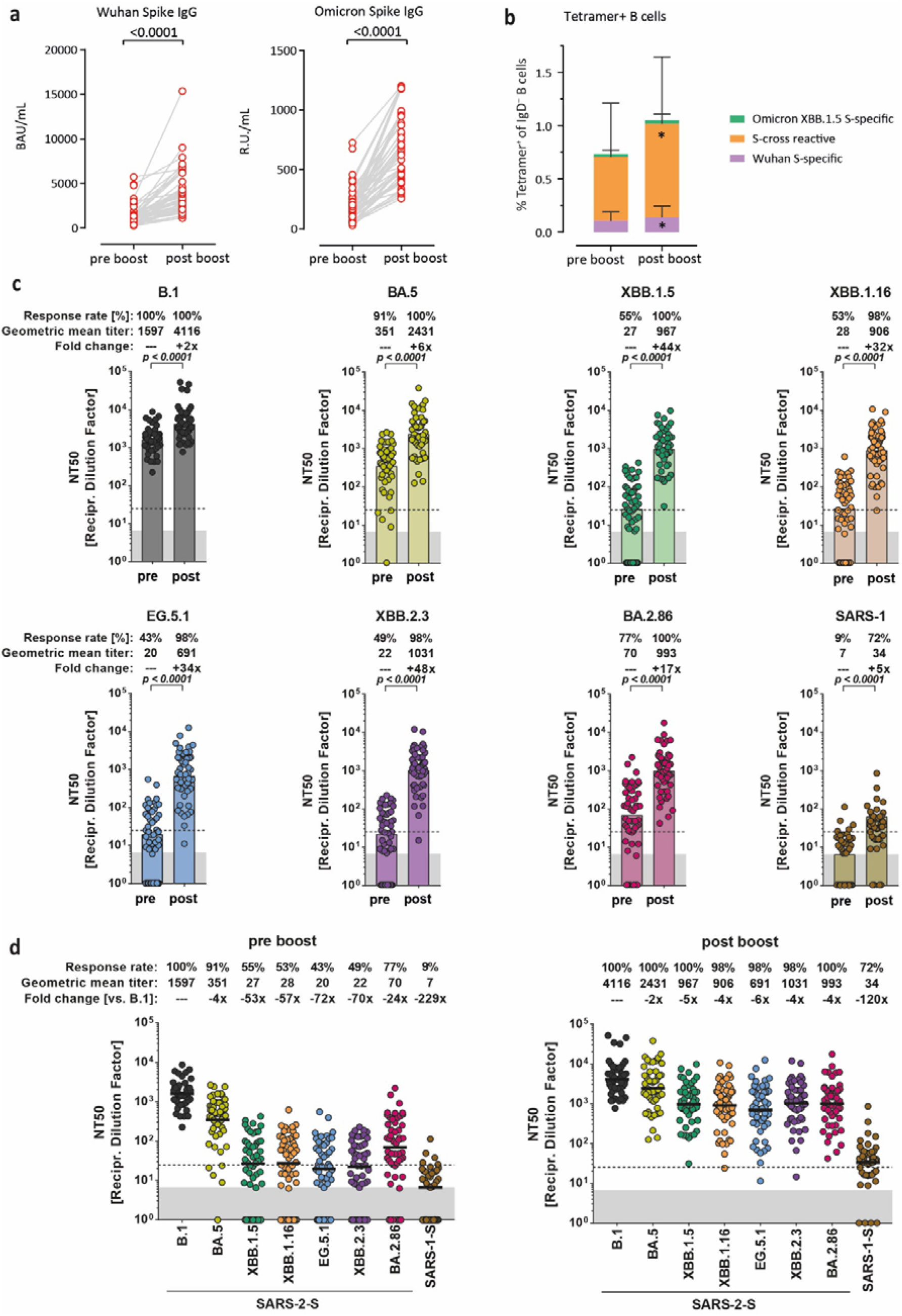
Humoral immune responses following BNT162b2 XBB.1.5 booster vaccination. **a.** Levels of Wuhan Spike-specific IgG and Omicron Spike-specific IgG in plasma taken before (pre-boost) or after booster vaccination (post-boost) with the BNT162b2 Omicron XBB.1.5 vaccine. Statistical significance was assessed by paired t-test. **b.** Frequencies of memory B cells binding Wuhan (Wu), Omicron XBB.1.5, or both (cross-reactive) RBDs before (pre-boost) or after booster vaccination (post-boost) with the BNT162b2 Omicron XBB.1.5 vaccine. Data are represented as mean+SD. **c.** Neutralization of pseudovirus particles (_pp_) bearing the indicated S proteins by donor-matched plasma (n=53) taken before (pre-booster) or after booster vaccination with the BNT162b2 Omicron XBB.1.5 vaccine (post-booster). Data represent geometric mean titers (GMT) from a single experiment, performed with four technical replicates. Information on response rates and median fold change in neutralization after booster vaccination are indicated above the graphs. Statistical significance was assessed by Mann Whitney test. Of note, for graphical reasons plasma samples yielding an NT50 value below 6.25 (limit of detection) were manually set at bottom of the axis. **d.** The data presented in panel c were re-grouped to compare differences in SARS-CoV-2 lineage-specific neutralization before and after vaccination. Information on response rates and median fold change in neutralization compared to B.1_pp_ are indicated above the graphs. See Extended Data Fig. 5 and 6 for individual neutralization data.

We next compared memory B cell (MBC) populations by measuring the frequency of Wu RBD-binding, XBB 1.5 RBD-binding, and both Wu RBD and XBB 1.5 RBD-binding cross-reactive MBCs by flow cytometry (Extended Data Fig. 3). For this, we used tetramerized recombinant spike proteins from Wu and Omicron XBB.1.5 variants to identify MBCs carrying B cell receptors binding to either one or both spike proteins^6^. As expected, pre-vaccination cross-reactive MBCs (mean frequency 0.6129 %) were the most prominent population compared to only Wu RBD-binding or XBB 1.5 RBD-binding MBC populations (mean frequency 0.1063 % and 0.0296 %, respectively). Post vaccination, the total amount of RBD-binding MBCs expanded significantly in line with the raise in anti-S IgG (Figure 1b). Cross-reactive MBCs also expanded significantly and continued to be the dominating MBC population producing anti-S IgG, while there was no change in the proportion of exclusively XBB 1.5 RBD-binding MBCs (Figure 1b, Extended Data Fig. 4) and little changes in only Wu RBD-binding MBCs (mean frequencies: 0.8786%, 0.0355% and 0.1367%). Overall, we observed a 40.3 % increase in RBD-binding MBCs, of which 88.0% was caused by the raise in cross-reactive MBCs. Although preliminary, these initial data suggest cross-reactive RBD-directed MBCs remained dominant even after multiple exposures to Omicron spikes including monovalent Omicron XBB.1.5 vaccination and underscore the role of persistent immune imprinting^2^.

We then analyzed neutralization using a pseudovirus particle (_pp_) neutralization assay (pVNT)^7^ including S proteins of seven SARS-CoV-2 lineages (Figure 1c and d, Extended Data Fig. 5 and 6) and SARS-CoV-1. We observed a 100% response rate at baseline for B.1_pp_, 91% for BA.5_pp_, 55% for XBB.1.5_pp_, 53% for XBB.1.16_pp_, 43% for EG.5.1_pp_, 49% for XBB.2.3_pp_, 77% for BA.2.86_pp_, and 9% for SARS-1_pp_ (Figure 1c). Particles bearing XBB sublineage S proteins were generally less efficiently neutralized compared to B.1_pp_ (median reduction, 53 to 72-fold), while SARS-1_pp_ neutralization was least efficient (229-fold reduction; Figure 1d). There were no major differences between neutralization of XBB.1.5_pp_, XBB.1.16_pp_ and XBB.2.3_pp_, as well as EG.5.1_pp_, while BA.2.86_pp_ evaded neutralization less efficiently. After vaccination, the response rates increased for all groups except B.1, which remained at 100% (Figure 1c and d). Importantly, a significant increase in neutralization for contemporary SARS-CoV-2 lineages was observed, with neutralization of XBB.1.5_pp_, XBB.1.16_pp_, EG.5.1_pp_, XBB.2.3_pp_, and BA.2.86_pp_ showing a median increase of 44, 32, 34, 48, and 17-fold, respectively. It has been shown, that monovalent mRNA-1273.815 (Spikevax XBB.1.5, Andusomeran, Moderna) vaccination similarly elicited a significant increase in neutralizing antibody titers, but to a lesser extend (7.5 to 14-fold) and was influenced by prior infection (https://doi.org/10.1101/2023.08.22.23293434). These differences may be due to the relatively low geometric mean titers before Omicron XBB.1.5 booster vaccination in our cohort as compared to the study assessing mRNA-1273.815^1^. Vaccinees in our cohort were younger, only 30% vs. 100% had received a previous dose of a bivalent vaccine and had a longer median interval from the last vaccine dose (14.6 vs. 8.2 months). Altogether, these data indicate that BNT162b2 Omicron XBB.1.5 booster vaccination induced a strong increase of S protein-specific antibodies that possess broad neutralizing activity against multiple XBB sublineages and BA.2.86.

In addition to humoral immune responses, we also analyzed frequencies of spike-specific T cells (Methods and Extended Data Fig. 7). The frequencies of interferon (IFN)-γ spike-reactive CD4^+^ and CD8^+^ T cells in blood samples collected before booster vaccination were relatively low compared to DMSO controls (Fig. 2a). After XBB.1.5 boosting, the frequencies for Wu and XBB.1.5 spike-responding IFN-γ-producing CD4 T cells increased significantly (p=0.0267 and p<0.0001, respectively). We observed similar significant increases (p=0.0225) for XBB.1.5 spike-responding CD8^+^ T cells (Fig. 2b). This is in line with evidence that memory CD8+ T cells elicited by mRNA vaccines recognize diverse spike epitopes ^8^. Application of the XBB.1.5 booster did not further expand Wu or XBB.1.5 responding spike-specific CD4^+^ and CD8^+^ T cells producing IFN-γ and/or tumor necrosis factor (TNF)-α (Extended Material Fig. 8). Finally, a significant increase in spike-responding IFN-γ-producing T cells after the booster vaccine was confirmed by IFN-γ measurement after SARS-CoV-2 Wu spike peptide stimulation of whole blood (Fig 2c). Whilst the mean IFN-γ concentrations doubled, resulting T cell responses were only somewhat above mean IFN-γ measurements obtained after the third vaccination^5^.

**Figure 2:**
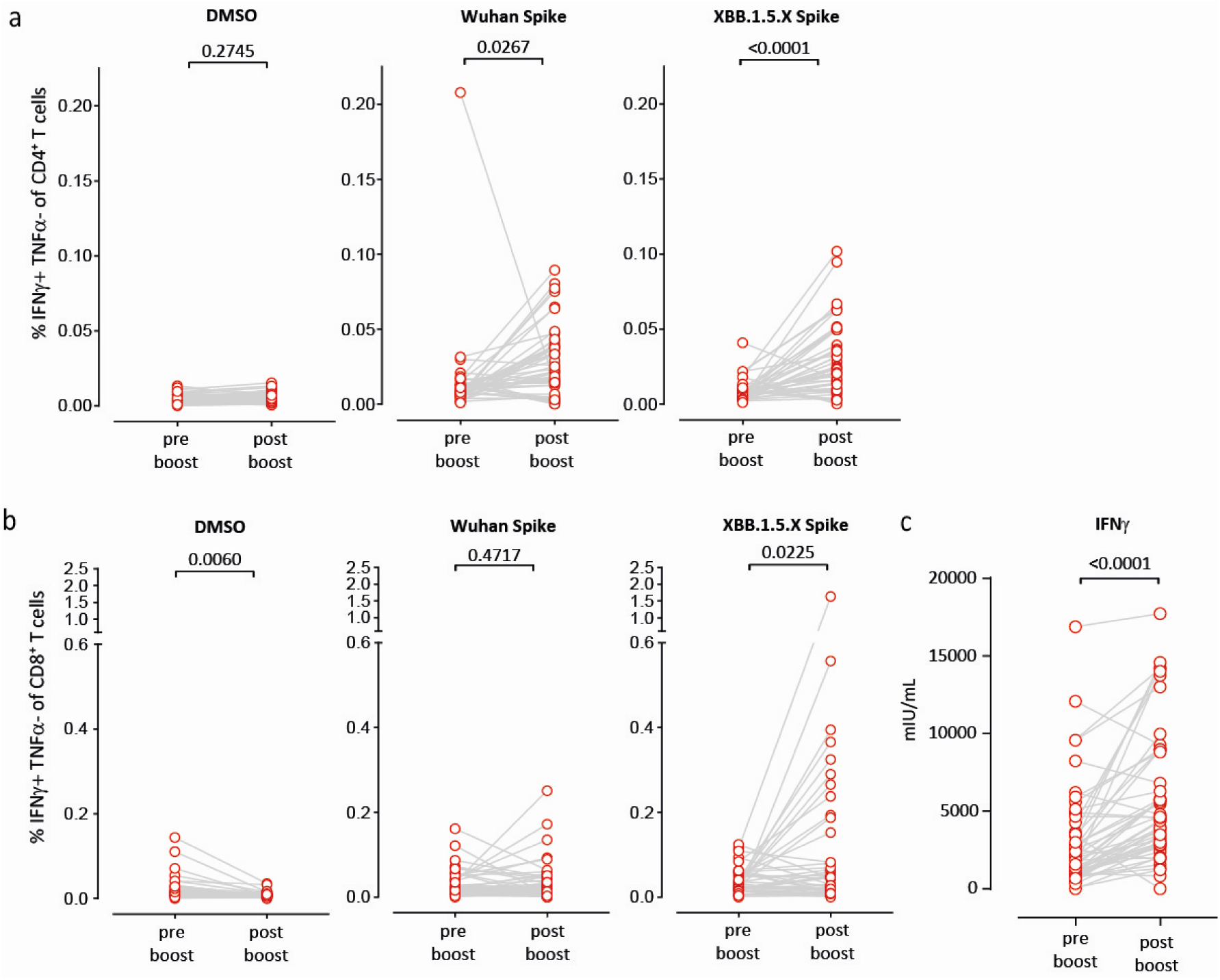
Cellular immune responses following BNT162b2 XBB.1.5 booster vaccination. **a, b.** Frequencies of IFN-γ^+^TNF-α− CD4+ **(a)** and CD8+ **(b)** T cells after exposure to DMSO only, Wuhan-Spike or XBB.1.5-derived peptides. T cells gated as shown in Extended Data Fig 7. Statistical significance was assessed by paired t-test. **c.** IFN-γ concentration in plasma after whole blood stimulation with SARS-CoV-2 S1 domain for 20– 24Lh. Statistical significance was assessed by paired t-test.

Clinical trials showed that neutralizing antibodies correlated with protection against the ancestral SARS-CoV-2 viral strains^9,10^. Although Omicron subvariants efficiently evaded vaccine-induced neutralizing antibody responses, protection against severe disease remained robust^11^. Thus, immune responses other than humoral immunity contribute to protection against severe disease. T cells recognize SARS-CoV-2-infected cells and restrict viral replication after infection. Unlike neutralizing antibodies, T cell responses induced by the original vaccines were highly active against Omicron variants^12–14^. These data, corroborated by CD8+ T cell depletion experiments in macaques^15^, suggest that T cell responses induced by vaccination (and infection) are critical for long-term protection against severe disease caused by SARS-CoV-2 variants.

The BNT162b2 Omicron XBB.1.5 monovalent vaccination analyzed here strongly increased anti-S IgG in all vaccinated individuals tested and elicited potent neutralizing responses against Omicron variants of the XBB lineage, including XBB.1.5, XBB.1.16, and XBB.2.3. The vaccine also induced cross-neutralizing responses against the more recent Omicron variants EG.5.1 (Eris) and BA.2.86 (Pirola). EG.5.1 is categorized as global variant of interest and efficiently evades neutralizing antibodies^16^. BA.2.86, which is antigenically distinct from XBB.1.5 and previous Omicron variants, can evade XBB-induced and XBB-effective neutralizing antibodies targeting various epitopes^17^. In sum, our data suggest that XBB.1.5 containing mRNA vaccines most likely increase protection against currently circulating XBB subvariants and the emerging BA.2.86 variant.

Our study has some limitations. Although daily monitoring of a post-vaccination subgroup confirmed that anti-Spike (S) IgG and neutralizing antibodies plateaued at day 8 to 10 post Omicron XBB.1.5 vaccination, consistent with antibody kinetics described after the second BNT162b2 vaccination ^18^ or after other COVID-19 vaccinations^19^, our data are preliminary and humoral and cellular immunity could further strengthen over time. Especially the development of XBB.1.5-specific MBCs warrants further monitoring. Second, most of our vaccinees had previous SARS-CoV-2 Omicron infections, likely contributing to considerable S protein responsive T cell immunity already before XBB.1.5 booster vaccination. Neutralization of SARS-CoV-2 lineages was assessed by pVNT, which has been shown to serve as an adequate surrogate model for this purpose^20^. Nevertheless, our data formally await confirmation with clinical isolates.

## Contributions

Study design: G.M.N.B., A.D.-J.

Data collection: M.V.S., R.G.J., A.C., G.M.R., T.G., G.A., T.W., S.I.H., M.F., I.R., T.I., J.R., A.S., S.W., M.H., A.K.

Data curation: A.C.

Data analysis: R.G.J., M.V.S., S.I.H., M.H., T.W., G.M.N.B, A.D.-J., R.F.

Data interpretation: G.M.N.B, A.D.-J., M.H., S.P., R.F.

Writing: G.M.N.B., A.D.-J. with comments from all authors.

## Ethics committee approval

The CoCo Study (German Clinical Trial Register DRKS00021152) and the analysis conducted for this article were approved by the Internal Review Board of Hannover Medical School (institutional review board no. 8973_BO-K_2020, last amendment Sep 2023). All study participants gave written informed consent and received no compensation.

## Data Availability

The data generated in this study are provided as a Source Data files in conjunction to this manuscript. All requests for raw and analyzed data that underlie the results reported in this article will be reviewed within four weeks by the CoCo Study Team, Hannover Medical School to determine whether the request is subject to confidentiality and data protection obligations. Data that can be shared will be released via a material transfer agreement.

## Acknowledgements

G.M.N.B. and A.D.-J. acknowledge funding (Niedersächsisches Ministerium für Wissenschaft und Kultur; 14-76103-184, COFONI Network, project 4LZF23) and A.D.-J. acknowledge funding by European Social Fund (ZAM5-87006761). S.P. acknowledges funding by the EU project UNDINE (grant agreement number 101057100), the Ministry for Science and Culture of Lower Saxony (Niedersächsisches Ministerium für Wissenschaft und Kultur; 14-76103-184, COFONI Network, projects 7FF22, 6FF22, 10FF22) and the German Research Foundation (Deutsche Forschungsgemeinschaft, DFG; PO 716/11-1). T.W. acknowledges funded by the DFG under Germany’s Excellence Strategy EXC 2155 “RESIST” (Project 390874280). Research in the Förster laboratory gets supported by the German Center for Infection Research TTU 01.938 (grant no 80018019238), by the German Center for Lung Research (grant 82DZL002B1), by the State of Lower Saxony (14-76103-184 CORONA-11/20), by DFG under Germany’s Excellence Strategy EXC 2155 “RESIST” (Project ID39087428), SFB 900/3 (Project B1, 158989968) and FO334/7-1 (all to R.F.).

We thank the CoCo Study participants for their support and the entire CoCo study team for help. We would like to thank Annika Heidemann, Janine Topal, Kerstin Sträche, Birgit Heinisch, Andrea Stölting, Emily Winter, Christine Jänke, Linus Risser, Marc Silchmüller, Sarah Long, Anna Zychlinsky Scharff, Nele Stein, Frank Müller, Sabine Buyny, Katja Kniesch, Svetlana Piter, Luise Graichen, Anna-Sophie Moldenhauer, and Eike Kreitz for technical and logistical support.

## Extended Data

**Extended Data Fig. 1.**
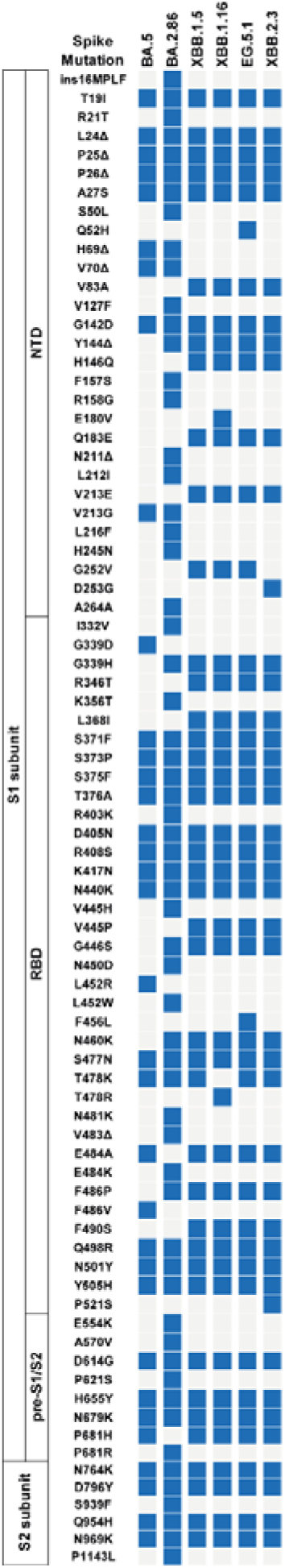
Overview on SARS-CoV-2 lineage-specific spike protein mutations.

**Extended Data Table 1.**
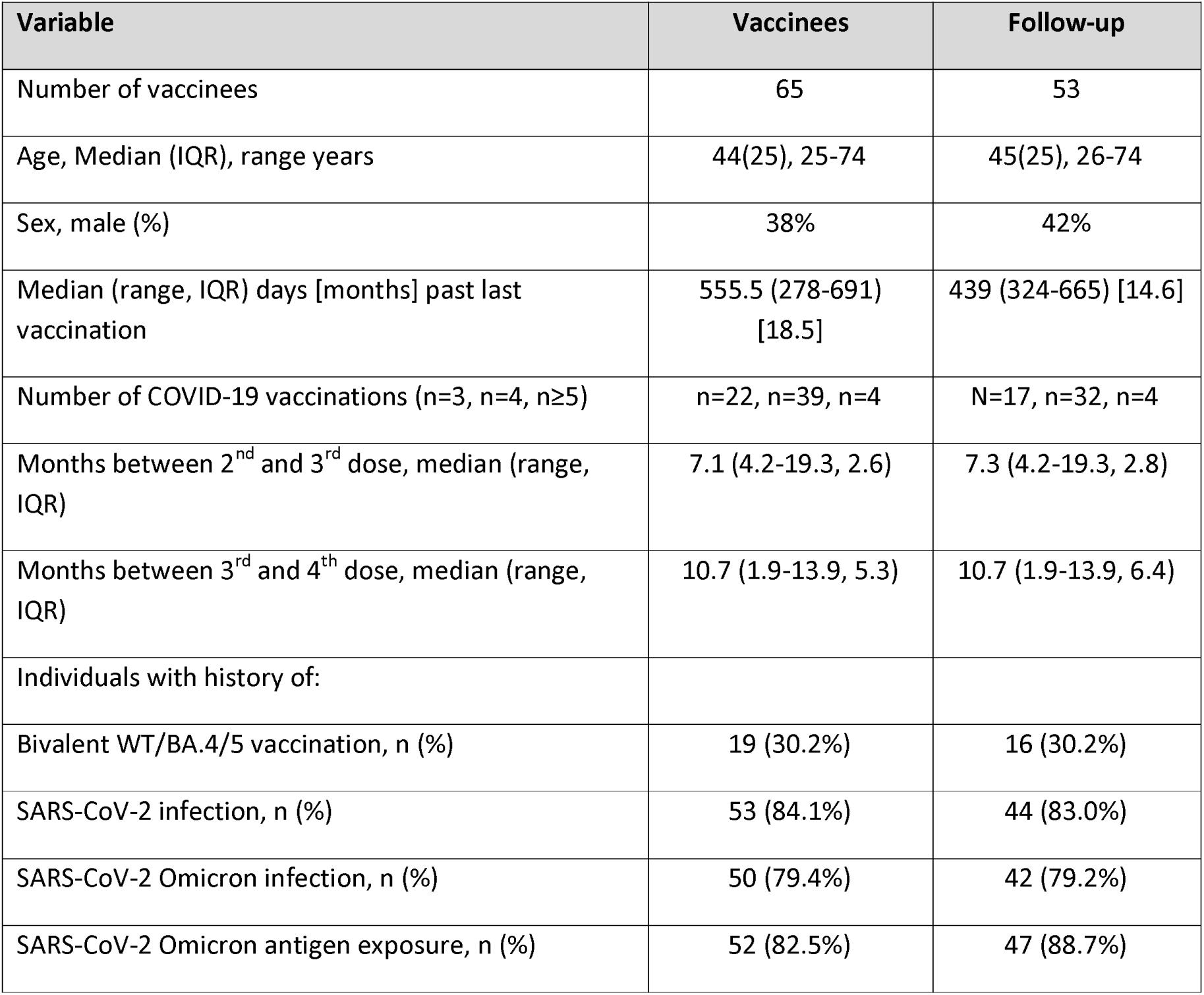

**Extended Data Fig. 2.**
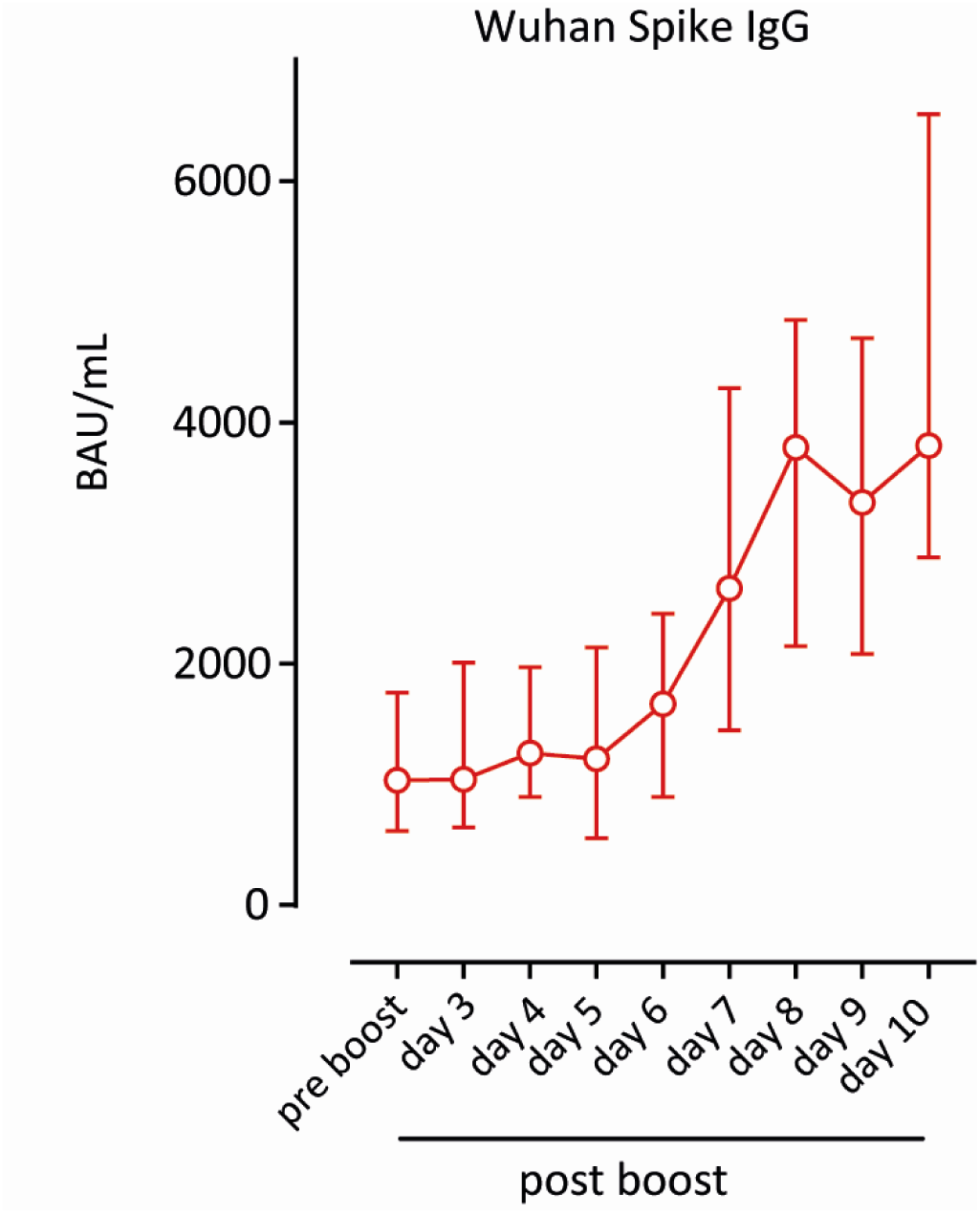
Longitudinal anti-S IgG measurements after vaccination in a sub group. Data depicted at day 3 to 8 include a sub group of up to 25 vaccinees. Results shown at pre boost and day 9 and 10 represent anti-S IgG of the entire cohort. Results are shown as median with interquartile range.

**Extended Data Fig. 3.**
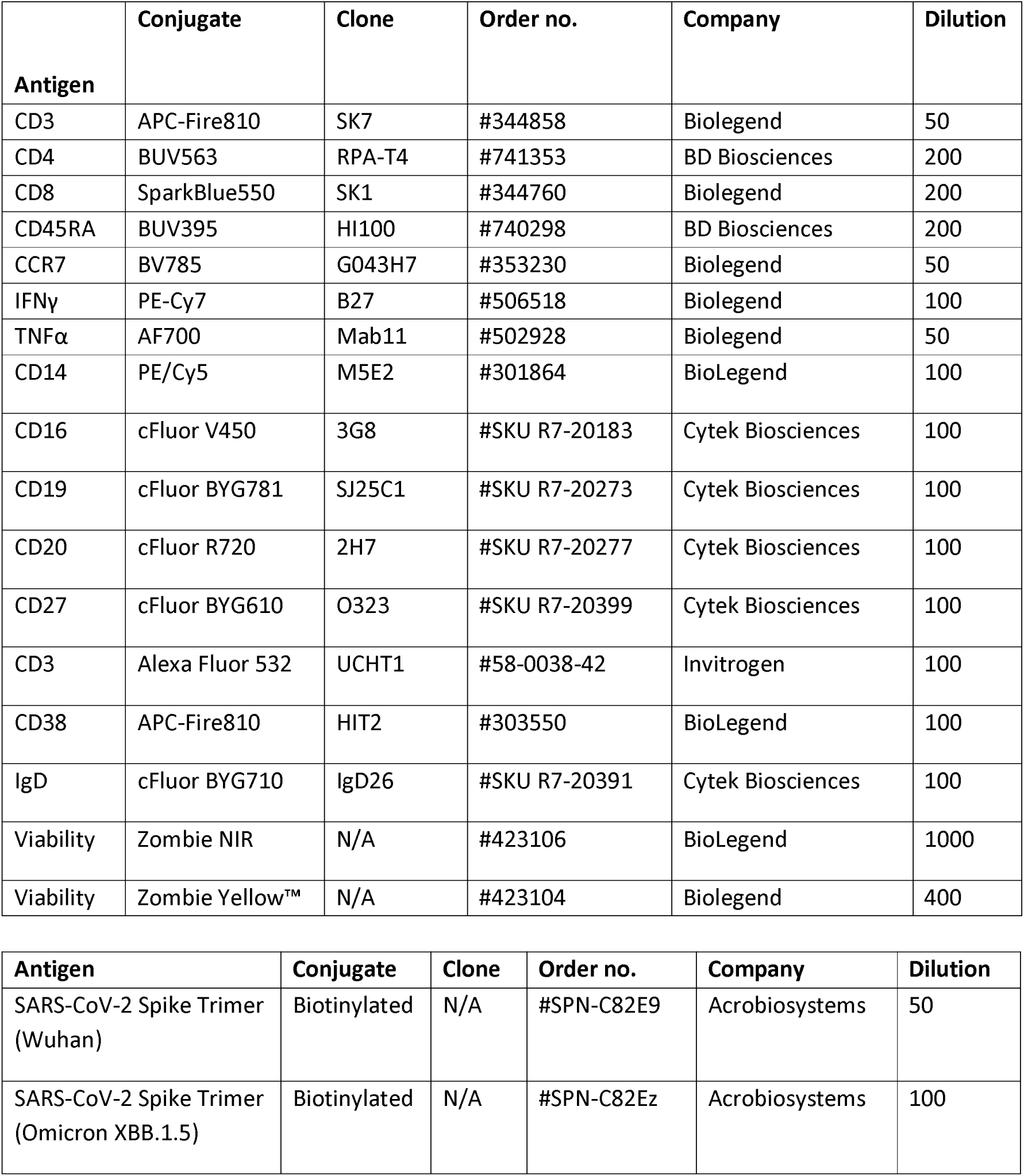
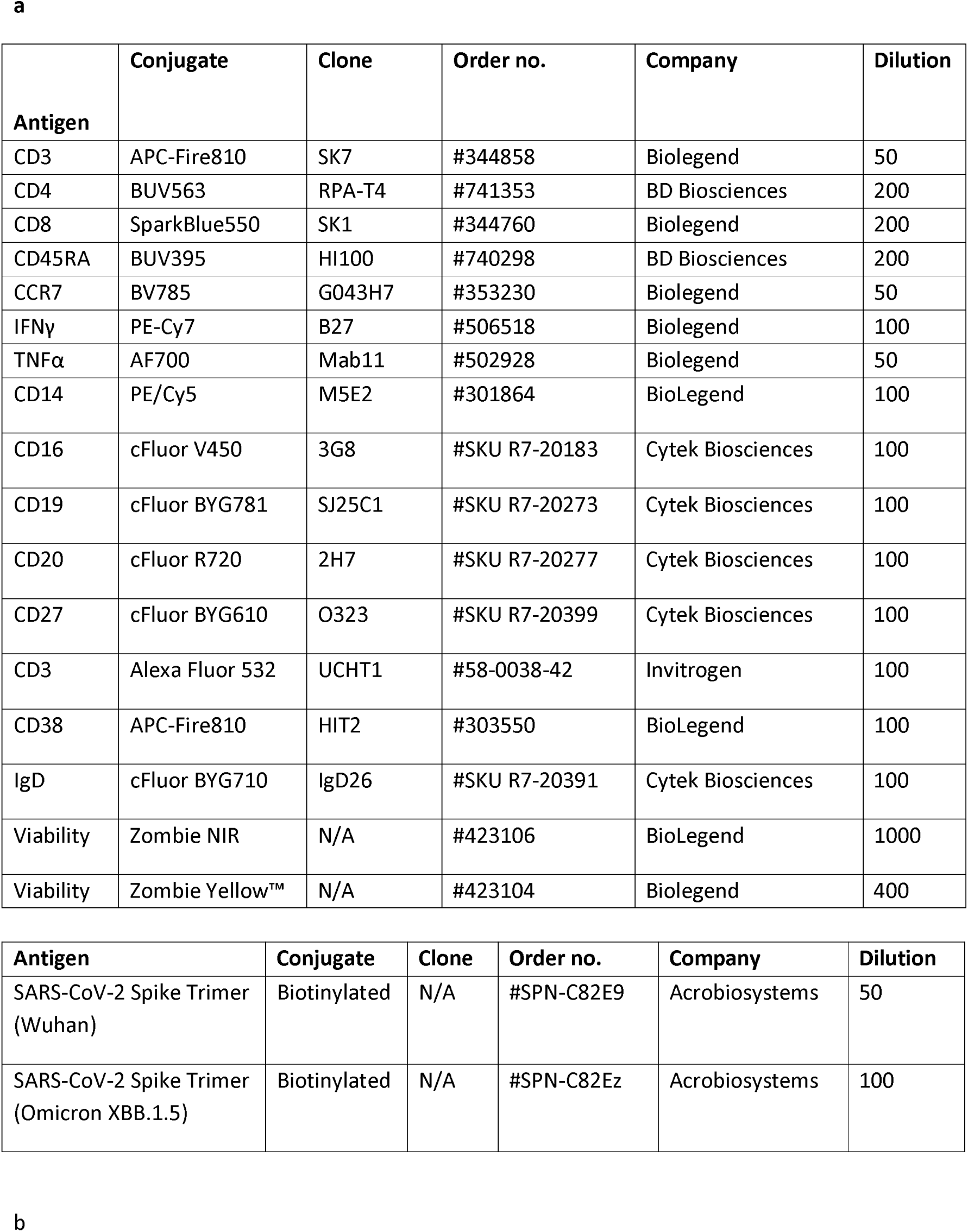
Antibody panel (a) and gating strategy (b) for SARS-CoV-2-S specific IgD^-^ cell populations in PBMCs. Tetramerized recombinant spike proteins from Wuhan (Tet Wuhan) and Omicron XBB.1.5 (Tet XBB.1.5) spike variants to identify MBCs carrying B cell receptors binding to either one or both spike proteins (Tet Wuhan/XBB.1.5). Pseudocolor plots show a representative data from a non-vaccinated individual.

**Extended Data Fig. 4.**
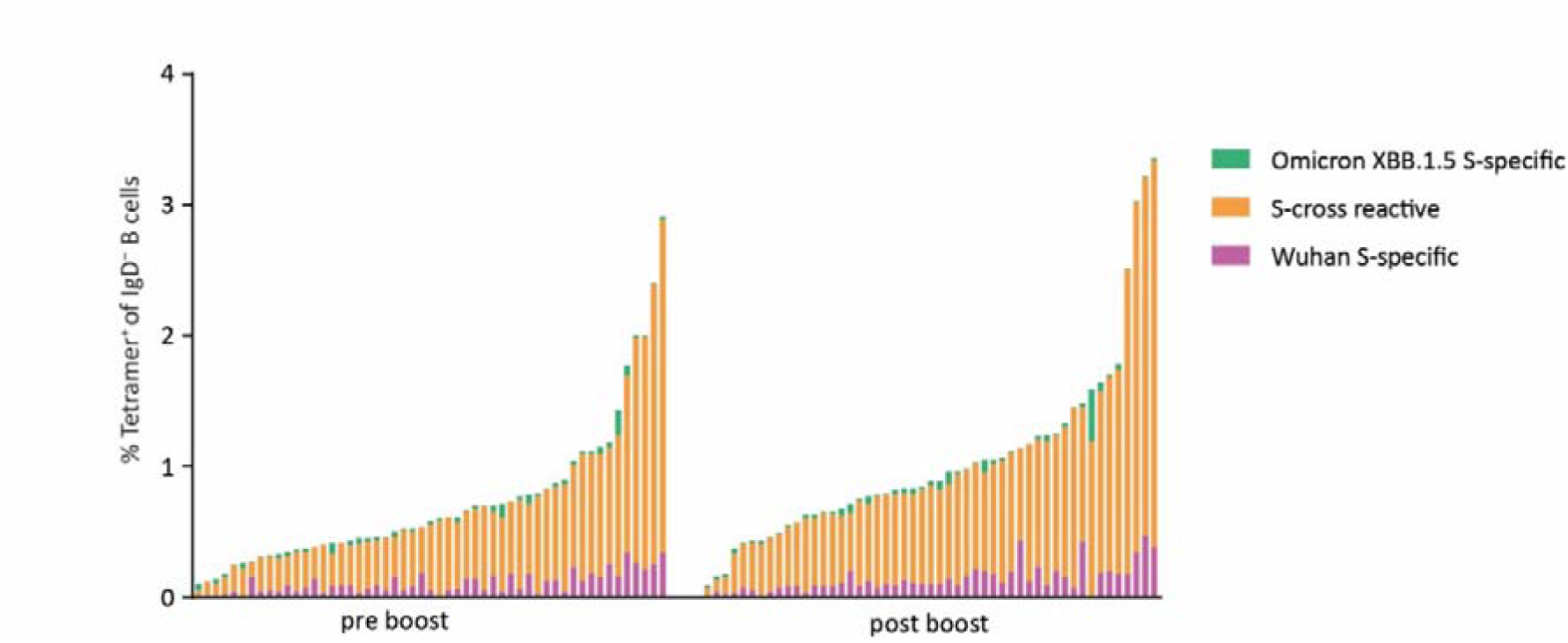
Individual data of changes in SARS-CoV-2-S specific IgD cell populations in PBMCs before and post Omicron XBB.1.5 booster. Tetramerized recombinant spike proteins from Wuhan and Omicron XBB.1.5 spike variants to identify MBCs carrying B cell receptors binding to either one (Omicron XBB.1.5 or Wuhan S-specific) or both spike proteins (S-cross reactive).

**Extended Data Fig. 5.**
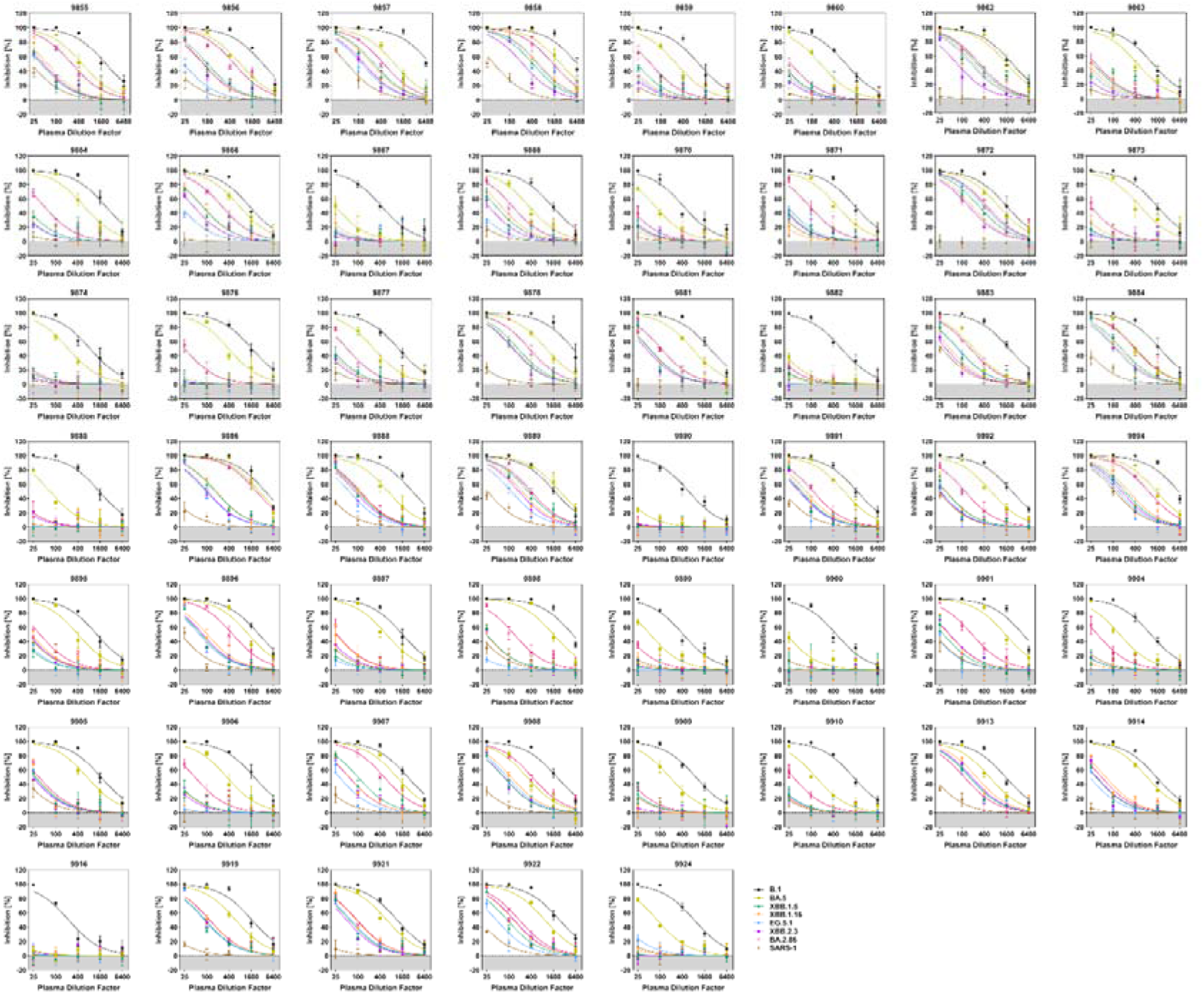
Individual neutralization data for pre-booster plasma.

**Extended Data Fig. 6.**
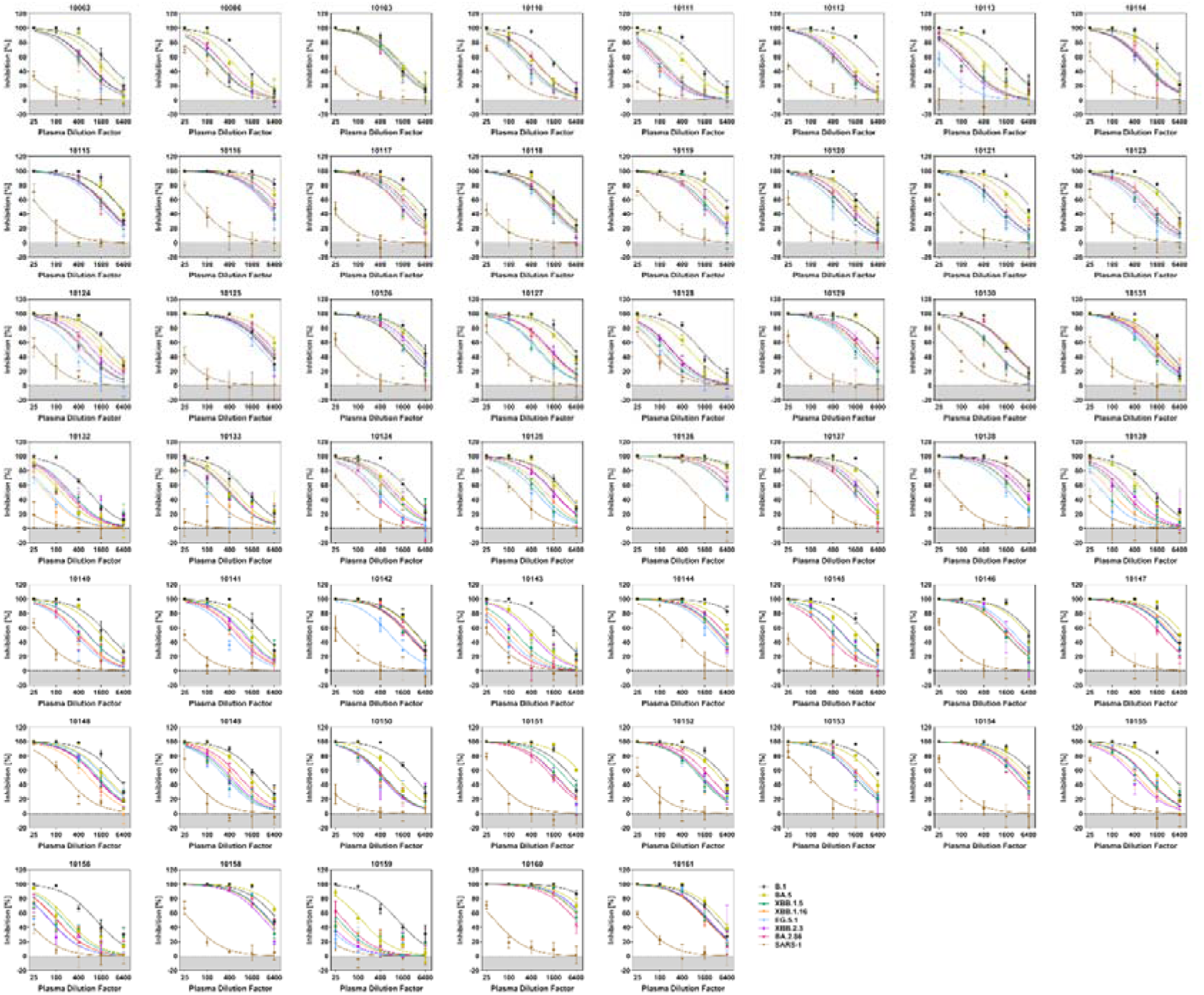
Individual neutralization data for post-booster plasma.

**Extended Data Fig. 7.**
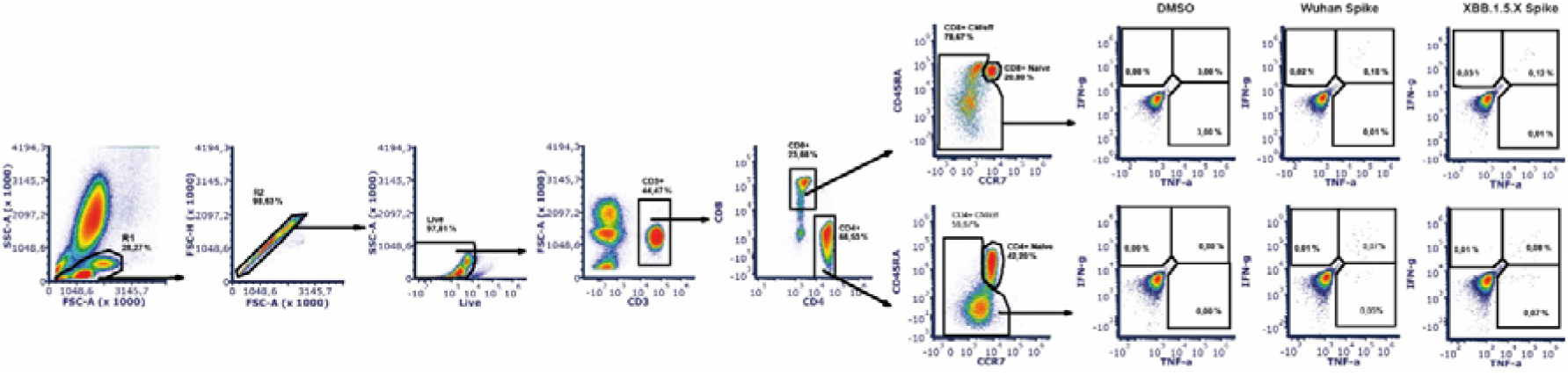
Gating strategy used for detection of cytokine-producing CD4 and CD8 T cells. We assessed cells after ex vivo re-stimulation with DMSO or Wuhan or XBB.1.5. Omicron Spike Peptide Pool for 12–16 hours.

**Extended Data Fig. 8.**
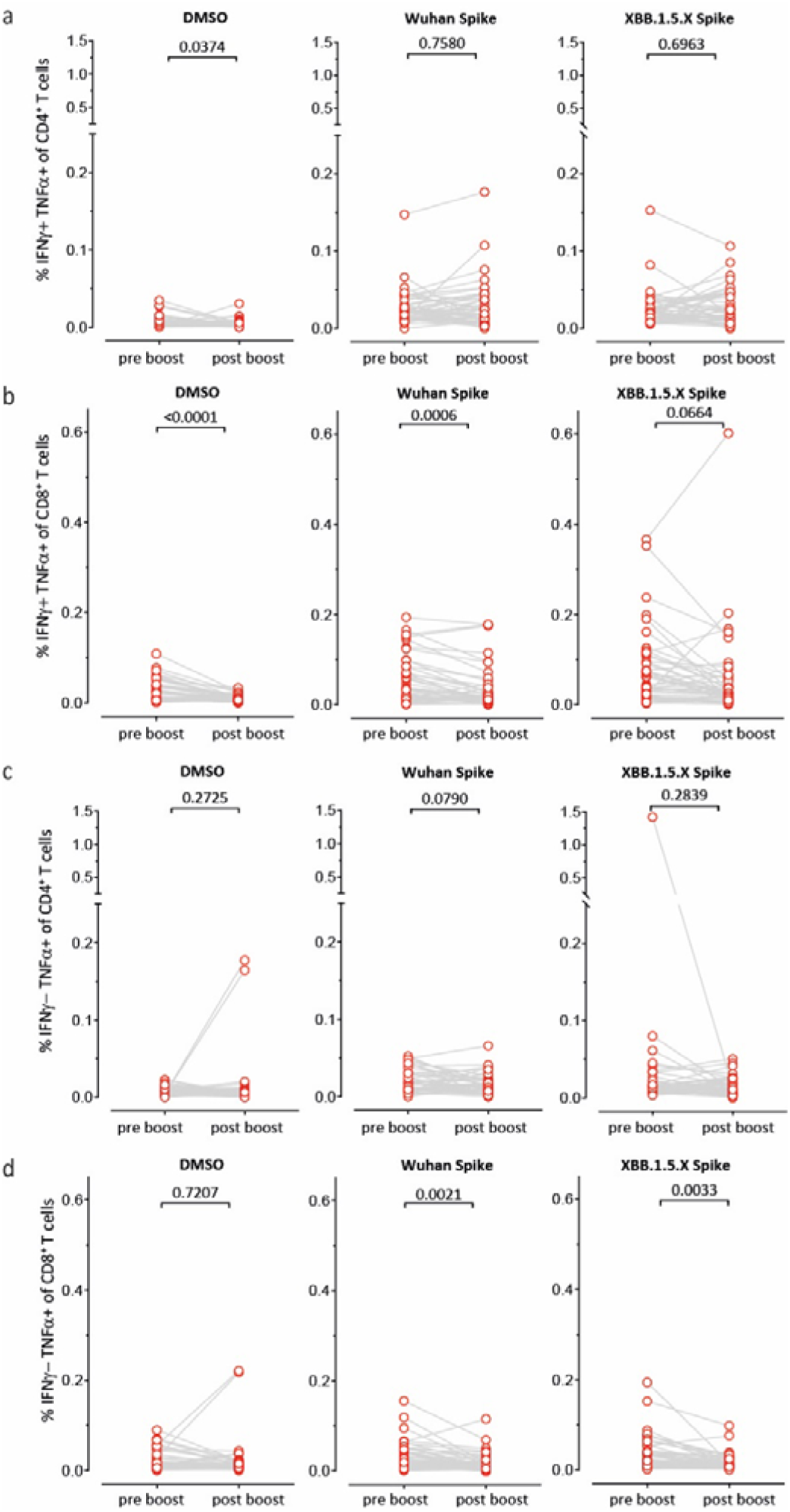
Composition of total cytokine-producing CD4+ and CD8+ T cells by the different subgroups. The figure depicts IFN-γ+TNF-α+ (a + b), and IFN-γ−TNF-α+ T cells (c + d) after restimulation with peptide pools derived from Wuhan S or Omicron XBB.1.5.

## Methods

### Participants

Participants in this analysis were 65 individuals vaccinated with BNT162b2 Omicron XBB.1.5 from the COVID-19 Contact (CoCo) Study (German Clinical Trial Registry, DRKS00021152), an ongoing, prospective, observational study monitoring anti-SARS-CoV-2 immunoglobulin G and immune responses in healthcare professionals at Hannover Medical School. In September 2023 study participants were offered COVID-19 vaccination with BNT162b2 Omicron XBB.1.5/raxtozinameran as part of the national vaccination campaign. Booster vaccination with BNT126b XBB 1.5 was coordinated independently and according to vaccine availability.

We estimated that a sample size of 42 would be sufficient to detect a clinically meaningful difference within the group, assuming that spike protein-specific IgG levels double from pre-vaccination (mean 822 RU/ml, standard deviation 747). This estimation was based on IgG measurements in a convenience sample of 24 persons from the CoCo cohort in August 2023 and on a one-tailed paired t-test of differences between means, with a 95% power and a 1% level of significance. Taking into account the above calculations and an expected loss-to-follow-up rate of 35%, a sample size of 65 vaccinated persons will be sufficient. The power calculation was performed using G*Power, Version 3.1.9.6.

Therefore, out of n=1,387 CoCo study participants, n=65 study participants that were vaccinated with BNT162b2 Omicron XBB.1.5 were invited to donate blood before and after BNT162b2 XBB 1.5 booster vaccination. To ensure the adequate timing of blood draws, a sub group of n=25 persons was followed with an increased frequency to monitor the increase of IgG up to the predefined endpoint of doubling the anti-spike (S) IgG antibodies and reaching a plateau. On day 7, this endpoint was reached, prompting participants to donate blood from day 8 to day 10. Median anti-S IgG antibodies did not show a significant increase from day 8 to 10 (see Extended Data Fig. 2). A total of n=53 vaccinees completed blood collection at a mean of 9.4 days post vaccination. Those who donated blood were not different from the entire group in terms of anti -S IgG antibodies at baseline, gender, age, and contacts with Omicron antigens. Data collection including questionnaires and lab assessment was done in Excel 2016.

No individual developed anti-SARS-CoV-2 NCP IgG after vaccination or reported a SARS-CoV-2 infection and was therefore excluded from follow-up analysis. Demographics (sex and age) and infection and vaccination history, respectively, are depicted in Extended Data Table 1. After blood collection, we separated plasma from EDTA or lithium heparin blood (S-Monovette, Sarstedt) and stored it at 4 °C for immediate use or at −80 °C until use. We used full blood or isolated PBMCs from whole blood samples by Ficoll gradient centrifugation and for stimulation with SARS-CoV-2 peptide pools.

### Serology

We measured SARS-CoV-2 IgG by quantitative ELISA (anti-SARS-CoV-2 S1 Spike protein domain/receptor binding domain IgG SARS-CoV-2-QuantiVac, EI 2606-9601-10G, and S1 Spike protein domain/receptor binding domain IgG SARS-CoV-2 of Omicron, EI 2606-9601-30 G, both EUROIMMUN, Lübeck, Germany) according to the manufacturer’s instructions (dilution up to 1:4000). We used anti-S1 concentrations expressed as RU/mL as assessed from a calibration curve with values above 11 RU/mL defined as positive^1^. We provide results obtained with the QuantiVac ELISA in binding antibody units (BAU/mL) converted by multiplying RU/mL by 3.2. We performed anti SARS-CoV-2 nucleocapsid (NCP) IgG measurements according to the manufacturer’s instructions (EUROIMMUN, Lübeck, Germany). We used an AESKU.READER (AESKU.GROUP, Wendelsheim, Germany) and the Gen5 2.01 Software for analysis.

### Production of pseudovirus particles and pseudovirus neutralization test (pVNT)

pVNTs were conducted at the Infection Biology Unit of the German Primate Center in Göttingen following a previously published protocol^2^ with minor modifications and the following S proteins were analyzed: B.1^3^, BA.5^4^, BA.2.86 (generated by Gibson assembly based on hCoV-19/South Africa/NICD-N25677/2021, EPI_ISL_8801154), XBB.1.5^5^, XBB.1.16^6^, EG.5.1^7^, XBB.2.3^7^, SARS-1-S^8^. In order to produce pseudovirus particles bearing S proteins of different SARS-CoV-2 lineages, 293T cells expressing the desired S protein upon transfection were inoculated with VSV*DG-FLuc^9^, a replication-deficient VSV vector that encodes for enhanced green fluorescent protein and firefly luciferase (FLuc) instead of VSV-G protein (kindly provided by Gert Zimmer, Institute of Virology and Immunology, Mittelhäusern, Switzerland). After 1 h of incubation at 37 °C, cells were washed with PBS and further incubated with medium containing anti-VSV-G antibody (culture supernatant from I1-hybridoma cells; ATCC no. CRL-2700). Pseudovirus particles were harvested at 16-18 h postinoculation. For this, the culture medium was collected and centrifuged (4,000 x g, 10 min). Subsequently, clarified supernatants were aliquoted and stored at −80 °C until further use.

pVNTs were performed in 96-well format using Vero76 cells (kindly provided by Andrea Maisner, Institute for Virology, Phillips University Marburg) as target cells. All plasma samples were heat-inactivated (56 °C, 30 min) before analysis and serially diluted in culture medium. In brief, equal volumes of pseudovirus particles and serially diluted plasma sample (final dilution range 1:25 to 1:6,400) were incubated for 30 min at 37 °C and subsequently inoculated onto confluent Vero76 monolayers. Pseudovirus particles incubated with medium alone instead of plasma sample served as controls. At 16-18 h postinoculation, efficiency of S protein-driven cell entry was analyzed by measuring FLuc activity in cell lysates. For this, the supernatant was aspirated, and cells were lysed with PBS containing 0.5 % Tergitol (Carl Roth; 30 min). Next, cell lysates were transferred into white 96-well plates, mixed with FLuc substrate (Beetle-Juice, PJK), and luminescence was recorded using a Hidex Sense Microplate Reader Software (version 0.5.41.0). Neutralization efficiency was determined based on the relative inhibition of pseudovirus entry, with signals obtained from pseudovirus particles incubated in the absence of plasma serving as reference (= 0% inhibition). In addition, a non-linear regression model was used to calculate the neutralizing titer 50 (NT50), indicating the plasma dilution required for half-maximal inhibition. Of note, plasma samples that yielded NT50 values below 25 were considered as non-responders. Further, plasma samples that yielded NT50 values below 6.25 (limit of detection, LOD) were assigned an NT50 value of 3.125 (0.5 of LOD).

### Flow cytometric detection and analysis of SARS-CoV-2-specific B cells

For tetramer preparation, we used recombinant, biotinylated SARS-CoV-2 spike (Acro Biosystems; Wuhan—SPN-C82E9 and Omicron/XBB.1.5—SPN-C82Ez) in order to detect SARS-CoV-2-spike-specific B cells. We tetramerized recombinant spike Wuhan proteins with fluorescently labeled streptavidin/R-phycoerythrin conjugate (Cat# S21388, ThermoFisher) and recombinant spike Omicron/XBB.1.5 proteins fluorescently labeled with streptavidin/allophycocyanin (Cat# S868;

ThermoFisher) according to the National Institutes of Health protocol as previously reported [10] (https://tetramer.yerkes.emory.edu/support/protocols). Briefly, we added targeting final equal molarity with moderate streptavidin excess, streptavidin to the recombinant proteins at 10 steps every 10 min. We incubated fresh PBMCs samples in complete RPMI at 2×10^6^ cells/mL for 6h at 37°C in 5% CO_2_ as previously described^10^. Next, we washed and re-suspended cells in FACS buffer (PBS, 1mg/mL BSA, 1mmol/L EDTA) and stained cells with antibodies according to the gating strategy (Extended Data Fig. 3) for 20 min at room temperature (RT). Finally, we added tetramerized recombinant proteins against the Wuhan spike or Omicron XBB.1.5 spike and incubated cells for an additional 20 min at RT. After two more wash steps, we acquired samples on a spectral flow cytometer (Cytek Northern Lights) and analyzed data using SpectroFlo and/or FCS Express software.

### T cell re-stimulation assay

After isolation by Ficoll gradient centrifugation, using Leucosep tubes, PBMCs were resuspended in complete RPMI medium [RPMI 1640 (Gibco)] supplemented with 1 mM sodium pyruvate, 50 µM β-mercaptoethanol, 10% fetal bovine serum (FBS) (GE Healthcare Life Sciences), and 1% streptomycin– penicillin (all from Gibco) at a concentration of 1 × 10^7^ cells per mL. PBMCs were then stimulated with XBB.1.5.X Omicron full length SARS-CoV-2 (Spike Glycoprotein) Peptide Pool (peptides&elephants; no. LB02145) or SARS-CoV-2 (Spike Glycoprotein) Peptide Pool (peptides&elephants; no. LB01792). All peptide pools were prepared in complete RPMI with a final concentration of 10 µg/mL brefeldin A (Sigma-Aldrich). The concentration of each peptide in the final restimulation mixture for 5 x 10^6^ cells was 0.3 nmol (approximately 0.5 µg)/mL. As a negative control, PBMCs were stimulated with DMSO only as the solvent of the peptide pools. As an internal positive control, in each experiment, cells were stimulated with ionomycin (Invitrogen) and phorbol 12-myristate 13-acetate (Calbiochem) at a final concentration of 1,500 ng/mL and 50Lng/mL, respectively. For the stimulation, cells were incubated for 12–16 h (at 37°C, in 5% CO_2_). After harvesting, cells were resuspended in MACS buffer (PBS supplemented with 3% FBS and 2 mM EDTA) containing 10% mouse serum and incubated at 4°C for 15 min to avoid non-specific antibody binding. Next, without washing, an antibody mix of anti-CD3-APC-Fire810 (SK7; # 344858; Biolegend; 1:50), anti-CD4-BUV563 (RPA-T4; #741353; BD Biosciences; 1:200), anti-CD8-SparkBlue550 (SK1; #344760; Biolegend; 1:200), anti-CD45RA-BUV395 (HI100, #740298, BD Biosciences; 1:200), anti-CCR7-BV785 (G043H7; #353230; Biolegend; 1:50) and Zombie Yellow™ Fixable Viability Kit (#423104; Biolegend; 1:400) was added. After staining for 20 min at RT, cells were washed before they were fixed and permeabilized (#554714; BD Biosciences) according to the manufacturer’s protocol. Next, intracellular cytokines were stained using anti-IFNγ-PE-Cy7 (B27; #506518; Biolegend; 1:100) and anti-TNFα-AF700 (Mab11; #502928; Biolegend; 1:50) for 45 min on RT. After washing off excess antibody, samples were analyzed on a five-laser Cytek Aurora spectral flow cytometer (Cytek) (355 nm, 405 nm, 488 nm, 561 nm, and 640 nm). All flow cytometry data were acquired using SpectroFlo version 2.2.0 (Cytek) and analyzed with FCS Express 7 (Denovo).

### Quantification of IFN-γ release

0.5 mL full blood were stimulated with manufacturer’s selected parts of the SARS-CoV-2 S1 domain of the Spike Protein for a period of 20-24 h (ET 2606-3003, SARS-CoV-2 Interferon Gamma Release Assay, IGRA (EUROIMMUN, Lübeck, Germany). We carried out negative and positive controls according to the manufacturer’s instruction and measured IFN-γ using an ELISA (EQ 6841-9601, EUROIMMUN, Lübeck, Germany). For analysis, we used an AESKU.READER (AESKU.GROUP, Wendelsheim, Germany) and the Gen5 2.01 Software.

### Statistics

Statistical analysis was conducted using GraphPad Prism 8.4 or 9.0 (GraphPad Software, USA) and SPSS 20.0.0 (IBM SPSS Statistics, USA). Outliers were included in the analysis, and missing values were excluded pairwise. Mean (SD) was used for normally distributed data, while median (IQR) was used for non-normally distributed data. In normally distributed data, paired t-tests were performed to assess intra-individual differences, and independent t-tests for between group differences, in non-normally distributed data, Wilcoxon-Mann-Whitney-tests were used. Neutralization titers were transformed to geometric mean titers. Differences were considered significant if p < 0.05. For change in spike protein-specific IgG levels a p of <0.01 was considered significant.

### Ethics committee approval

The CoCo Study (German Clinical Trial Register DRKS00021152) was approved by the Internal Review Board of Hannover Medical School (institutional review board no. 8973_BO-K_2020, last amendment Sep 2023).

